# Peripheral Zone Enlargement: a newly described entity on MRI

**DOI:** 10.1101/2020.04.03.20052290

**Authors:** Neil F. Wasserman, Paari Murguan, Ben Spilseth, Gregory J. Metzger, Tina Sanghvi

**Affiliations:** Departments of Radiology, University of Minnesota, Minneapolis, MN; Laboratory Medicine, University of Minnesota, Minneapolis, MN

**Author notes:** Corresponding Author: Neil F. Wasserman, Department of Radiology, Univ. of Minnesota Medical School, Mayo Mail Code 292, 420 Delaware Street S.E., Minneapolis MN 55455. This study had IRB and Ethics Committee approval # 2871. Patient consent waved. The authors have no grants or disclosures to report.

**Keywords:** Prostate, Peripheral Zone, Enlargement, Measurement

## Abstract

**Purpose:** A retrospective study was performed to describe and characterize a previously unreported finding on T2-weighted magnetic resonance imaging (MRI) of peripheral zone enlargement (PZE) associated with total prostatic enlargement without necessary association with benign prostatic hyperplasia (BPH) or lower urinary tract symptoms.

**Methods:** T2-weighted MRI were reviewed from 2012-2018. Patients were referred for elevated serum prostatic specific antigen (PSA) levels, prostatic enlargement, or abnormal digital rectal examination (DRE) suggestive of adenocarcinoma. Enlargement of the thickness of the peripheral zone (PZ) was defined as a measurement of postero-lateral thickness (PLPZ) of ≥ 15.8 mm in the maximal transaxial plane. Endorectal coils were used in 2 patients. Microscopic pathology was described in 3 patients.

**Results:** There were 22 patients who met the criteria out of 2871 subjects (<1%). Mean age was 63 years, Mean PLPZ was 17.7 mm (CI=0.45, range, 15.8-21.3). mean total prostatic volume was 55.1 cc (range, 19.9-127.2 cc), mean transition zone volume was 20.6 cc (range, 2.7-71.9 cc), mean transition zone index (TZI) was 0.34 (range, 0.31-0.68), mean prostatic specific antigen (PSA) was 13.9 ng/mL (range, 0.28-144.7), mean body mass index was 28.9 (range 23.0-36.3).. Enlargement was described in 14 of 20 (70%). Pathological findings showed marked glandular distention and atrophy with interstitial edema and chronic inflammatory cells. Significance of these results is discussed.

## Introduction

The use of magnetic resonance imaging (MRI) has become the preferred modality to assess the prostate gland for many conditions presenting clinically to the urologist. It is useful in investigation of patients with elevated prostate specific antigen (PSA) to distinguish benign conditions such as benign prostatic hyperplasia (BPH) and prostatitis from cancer, and MRI is increasingly utilized in the diagnosis and staging of adenocarcinoma of the prostate [1, 2].

While imaging assessment of prostate size in patients with lower urinary tract symptoms (LUTS) may start with transrectal ultrasound (TRUS), details of lobar distribution may require MRI. These latter factors may determine whether surgery, minimally invasive techniques or medical management will provide the best outcome [3]. Imaging may also influence which class of medication, alpha blockers or 5-alpha reductase inhibitors, are likelier to result in optimal symptom relief [3].

Occasional patients presenting with LUTS and/or prostatic enlargement on digital rectal examination (DRE) display an apparent disproportionate thickness of the peripheral zone (PZ) relative to transition zone (TZ) size (Fig 1]. Normal parameters for PZ thickness have been recently reported to equal a mean of 10.0 mm and ≥15.8 mm maximum thickness representing 2SD above the mean [4].

**Fig 1.**
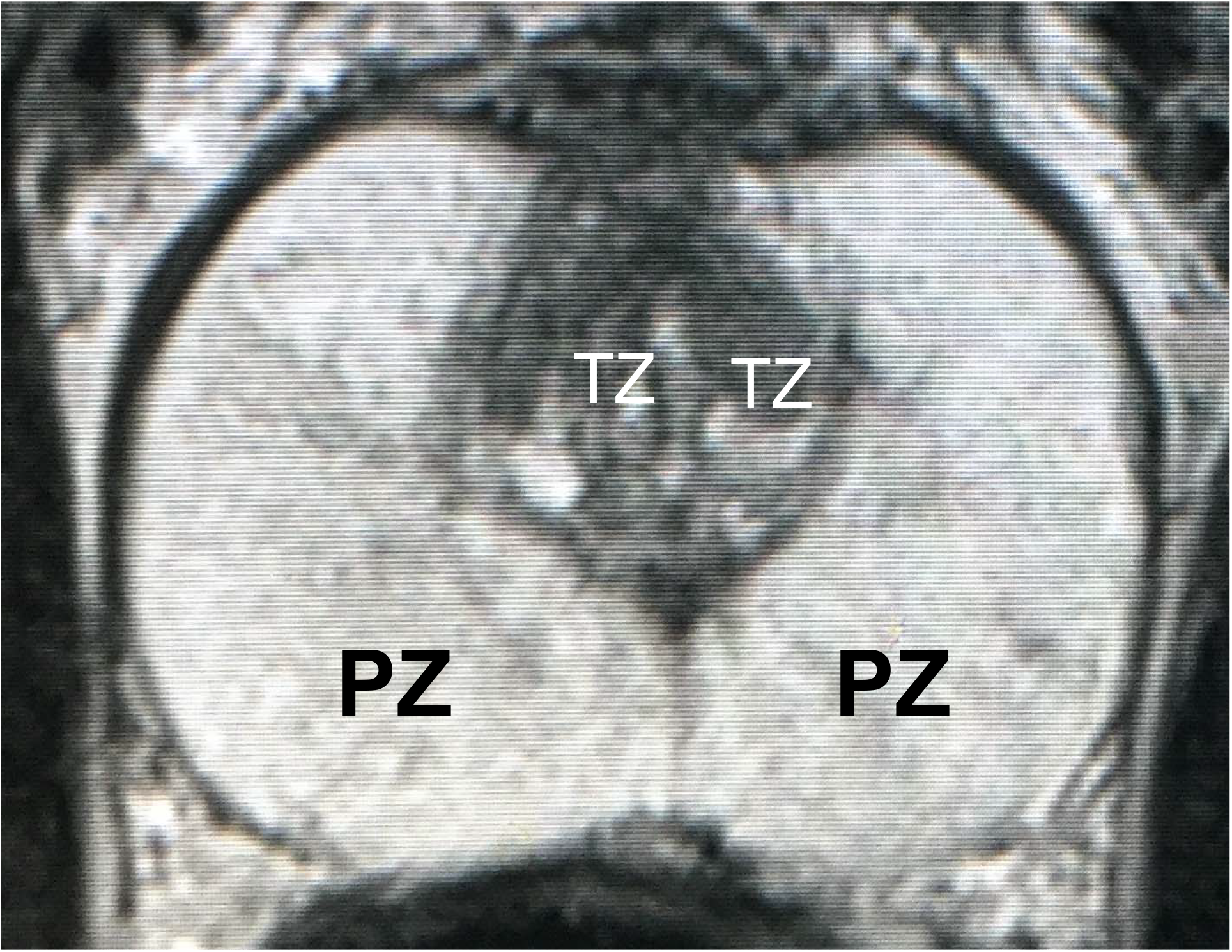
T2-weighted MRI at the maximal axial plane in 48-year-old with history acute prostatitis 3 years previously and currently with Mondor’s disease. Prostatic specific antigen is 3.6 µgm/mL and enlarged prostate on digital rectal examination. MRI measured total prostatic volume at 46 cc and interpreted as showing “BPH”. PZ=hyperintense peripheral zones, TZ=transition zones. Note disproportionately large PZ relative to TZ.

The purpose of this article is to report and characterize a previously unreported finding on T2-weighted MRI of PZ enlargement manifested by measurable thickening in the axial plane. This institutional review board (IRB) and Ethics Committee-approved study # 2871 retrospectively examined all MRI examinations at our institution over a period of 6 years searching for patients with abnormally thickened peripheral zones. We will show the results of this search, possible pathological cause of this finding and discuss its implications.

## Material and Methods

### Patient Population

We reviewed all MRI in our PACS and hospital electronic records over the period from 2012-2018 under approval of our IRB and Ethics committee #2871. Patient written permissions were waved for this retrospective study. We excluded MRI exams that were technically unsatisfactory because of large focal tumor distorting the thickness of the PZ or so extensive that thickness of the PZ could not be accurately measured. Other exclusions included previous prostate surgery, minimally invasive procedures, or pelvic radiation. The data set was left with 2871 individual patients, some of whom had more than one examination. These patients had been referred for MRI because of elevated PSA, abnormal DRE suggestive of malignancy, and/or prostatic enlargement.

### MRI Technique

Prostate MRI was performed using 3T for all patients (Magnetom Skyra or TrioTim; Siemens Healthcare; Erlangen Germany). The majority were performed using a pelvic phase array protocol without endorectal coil (ERC), and less that 10% were performed with Endorectal coil (Medrad, NJ, USA). The exact imaging parameters evolved during the study period, and at all times PI-RADS compliant technique was utilized. A typical protocol during the study period included T2W fast spin echo (FSE) imaging in the axial plane (TR/TE 3700/111 msec; NEX 3; 3mm slice thickness; no interslice gap; flip angle 160 deg; FOV 140 mm, matrix 320 × 256) and coronal plane (TR/TE 4030/100 msec; NEX 2; 3 mm slice thickness; no interslice gap; flip angle 122 deg; FOV 180 mm; matrix 320 × 256). T2W 3D SPACE images were obtained (TR/TE 1400/101 msec; flip angle 135 deg; 256 × 256 × 205 matrix, 180 mm FOV). T2W 3D SPACE images were reconstructed at 1 mm in the axial plane and also at 3mm in the, axial, sagittal, and coronal planes. Additionally, dynamic contrast enhanced T1-weighted (3 mm slice thickness; TR/TE 4.9/1.8 msec; 224 × 156 matrix, 250 mm FOV, temporal resolution <10 sec), and diffusion-weighted images at b values of 50, 800, and 2000 s/mm3 were performed. Endorectal coils were used in 2 patients.

### Measuring Techniques and Analysis

All images were reviewed, and PZs measured on PACS (Philips Intellispace v4.4 software) on high resolution monitors by a single board-certified radiologist with over 40 years of experience. The reader was blinded to the patient electronic medical record (EMR). Total prostate volumes (TPV) were measured using the biproximate prolate ellipsoid model (length × width × anteroposterior diameter × 0.52). Length measurement in the mid-sagittal plane used a line drawn at the vesico-prostatic junction (the vesico-prostatic line [VPL]) [5], and length measurement was made perpendicular to this line from the apex. Any prostatic tissue above the VPL was measured separately as a perpendicular from the proximal margin of the tissue to the VPL and added to the measurement below the VPL to derive total length of the prostate. This procedure had been previously demonstrated to have inter-observer reliability and validated against the traditional method of measuring length from the proximal prostatic margin to the apex [5]. The traditional prolate ellipsoid measurement boundaries and formula was used to calculate the TZ volume (TZV). Endorectal coils were used in 2 patients.

Out of the original patient set, 22 cases measured, posterolateral PZ thickness ≥15.8 mm. Peripheral zone measurements were independently made from T2 MRI images on this subset by a second board-certified diagnostic radiologist with 5-year experience using prostate volume metrics. The right and left PLPZ measurements were summed and averaged for each of the 2 observers. The final mean for each patient represented the summed and averages of both examiners.

The distribution of the PZ extends from the prostatic base to its apex. Measurements were made in the axial plane at the level of largest transverse diameter. Right and left posterolateral PZ (PLPZ) measurements were made from the estimated outer margin of the periurethral hyperplastic tissue to the inner boundary of the EPC following a perpendicular line defining the longest distance to the EPC [4](Fig. 2). The mean of the right and left PLPZ in millimeters (mm) was determined for each examiner, and the average of the two means recorded as the “composite” (pooled) PLPZ thickness [4].

**Fig 2.**
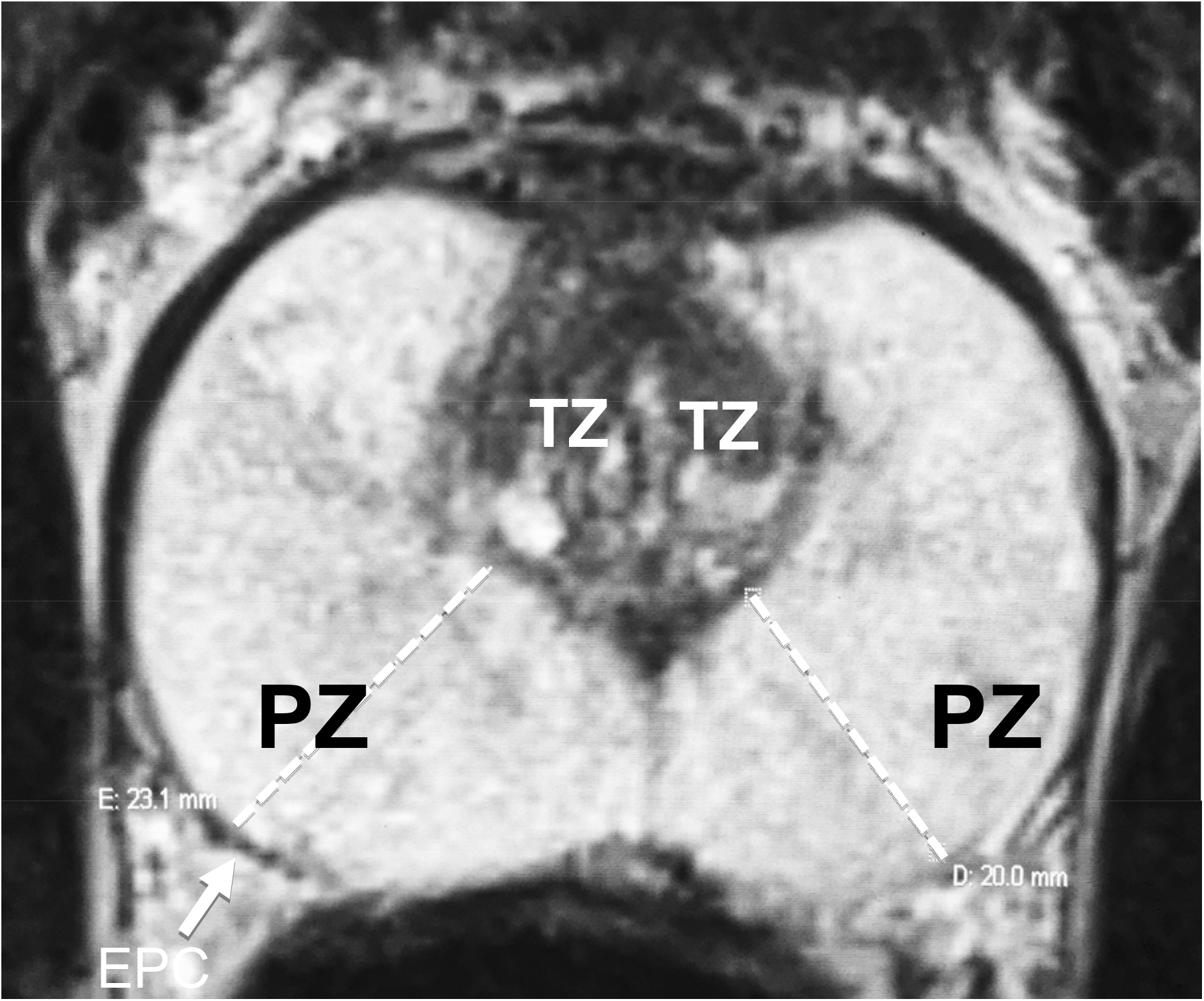
(Same patient as figure 1) T2-weighted MRI in maximal axial plane showing normal-sized transition zone (TZ) and enlarged peripheral zones. Right and left posterolateral peripheral zone measurements (dotted lines) measure mean of 21.5 mm from outer boundary of TZ to inner boundary of the external prostatic capsule (EPC).

In addition to measured variables, we analyzed calculated ones, particularly the transition zone index (TZI) representing the ratio of the TZ volume to TPV. PZ volume (PZV) was calculated by subtracting TZ volume from TPV [6]. Additional MRI observations were recorded. After all measurements were made by the 2 observers, the senior examiner reviewed the EMR and for clinical, laboratory and pathological data for correlation with MRI findings.

## Results

### PZE selection criteria

There were 22 patients who met the criteria for PZE out of 2871 subjects (<1%). Mean age was 63-years. Mean PLPZ was 17.7 mm (CI=0.45, range 15.8-21.3). Mean total prostatic volume was 55.1 cc (range, 19.9-127.2), mean transition zone volume was 20.6 cc (range, 2.7-71.9), mean transition zone index (TZI) was 0.34 (range, 0.31-0.68), mean prostatic specific antigen (PSA) was 13.9 ng/mL (range, 0.28-144.7), mean body mass index (BMI) was 28.9 (range 23.0-36.3). Median values were PLPZ= 16.2 mm, PTV=46.9 cc, TZV=18.3 cc, TZI= 0.34, and PSA=7.25 ng/mL). Enlargement was described in 14 of 20 (70%) in which digital rectal examination was recorded in the hospital record.

### Other Findings

There were 6 patients with type 2 diabetes mellitus, 4 with clinical history of prostatitis. American Urological Association Symptom Scores that were recorded (12) averaging 7.92 (range 0-23). DRE was described in the records of 20 patients and suggested enlargement in 14 (70%).

### MRI findings

The PZ is seen as a broad hyperintense band surrounding the darker mixed echogenic TZ. MRI findings were non-specific showing radiating dark bands extending through the PZ from the TZ toward the capsule in 9 cases, 5 of which were striking (Fig. 3). Five patients were PIRAD-2, 4 PIRADS-3, 1 PIRADS-4, the others unclassified. Lobar classifications of benign prostatic hyperplasia (BPH) [5] included 2 without BPH (PZ Type 0), 9 PZ, 2 PZ2, 8 PZ3, and 1 PZ5.

**Fig 3a,b.**
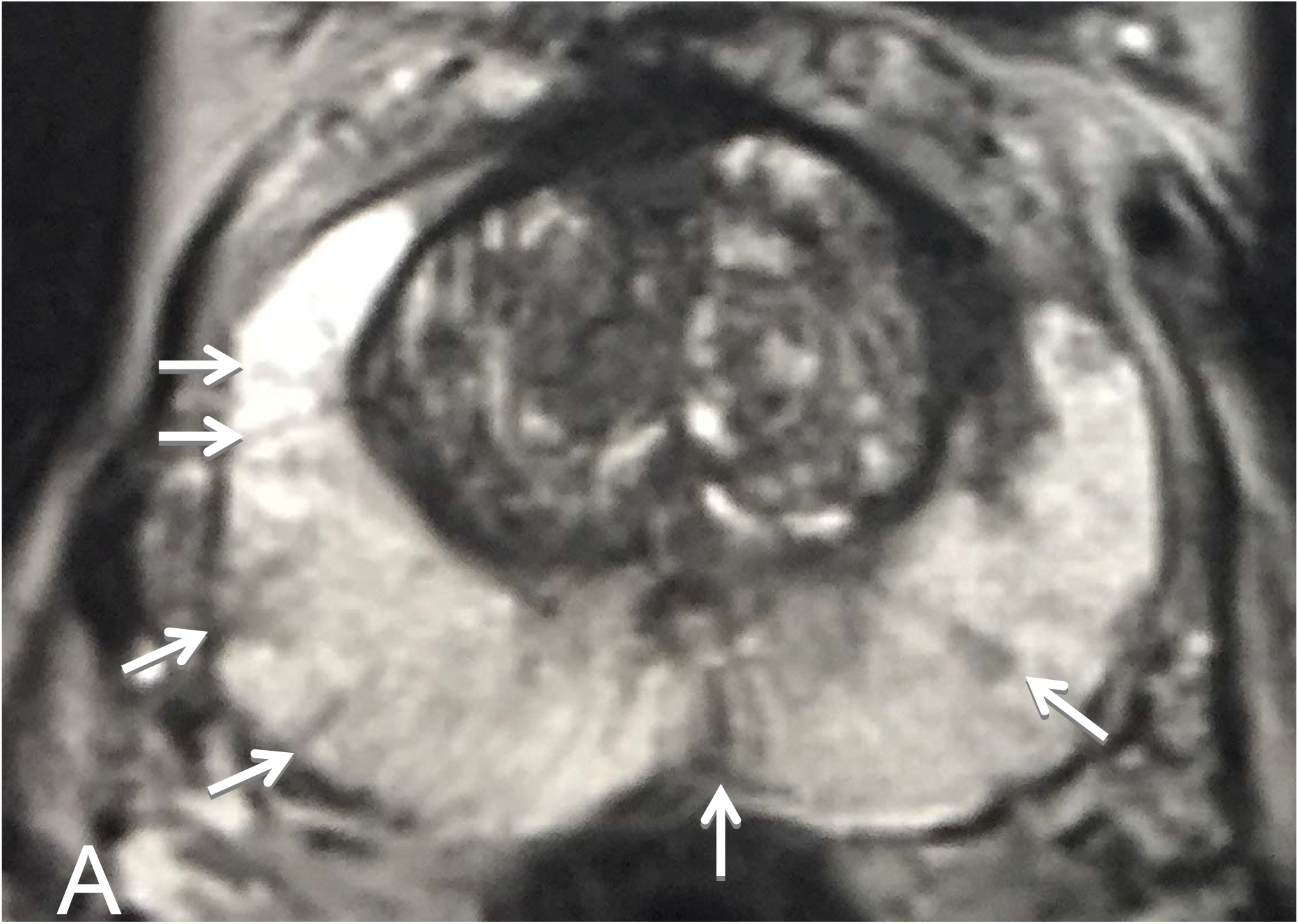
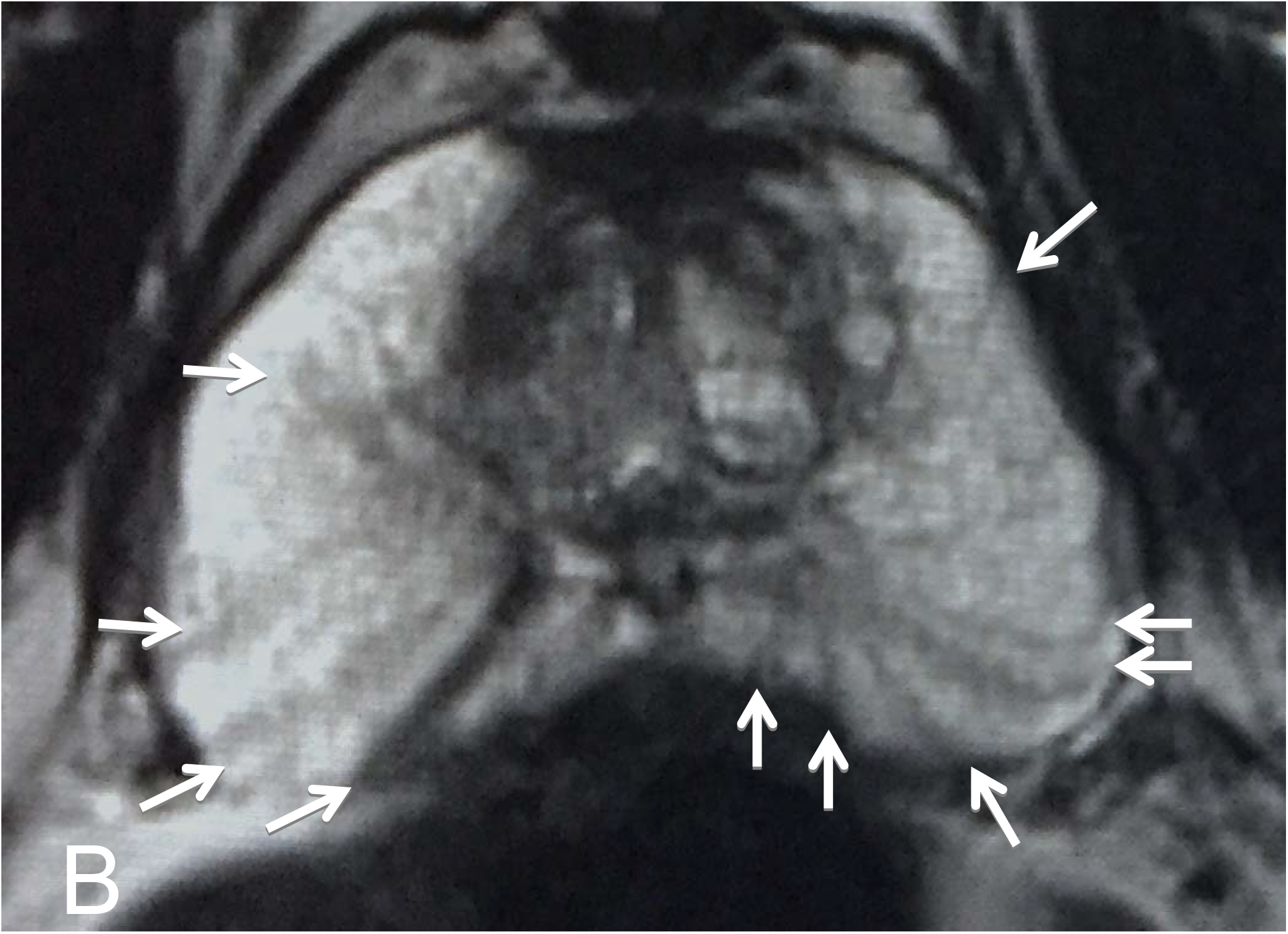
T-2 weighted MRI showing dark bands in peripheral zones (arrows), many of which extend to external prostatic capsule. Mean posterolateral peripheral zone thickness =18.7 mm in the maximal axial transverse plane.

### Pathological Findings

Total prostatectomy tissue was available in 3 of our 22 patients. Pathological findings showed marked PZ glandular distention and atrophy with interstitial edema and chronic inflammatory cells (Figs. 4,5). No ductal dilation between the glandular elements of the TZ and the verumontanum were observed. Transrectal ultrasound-guided biopsies were obtained in 17 patients and “negative for malignancy” in 8 (47%) with 1 showing “prostatitis” and positive for adenocarcinoma in 8 (47%).

**Fig 4.**
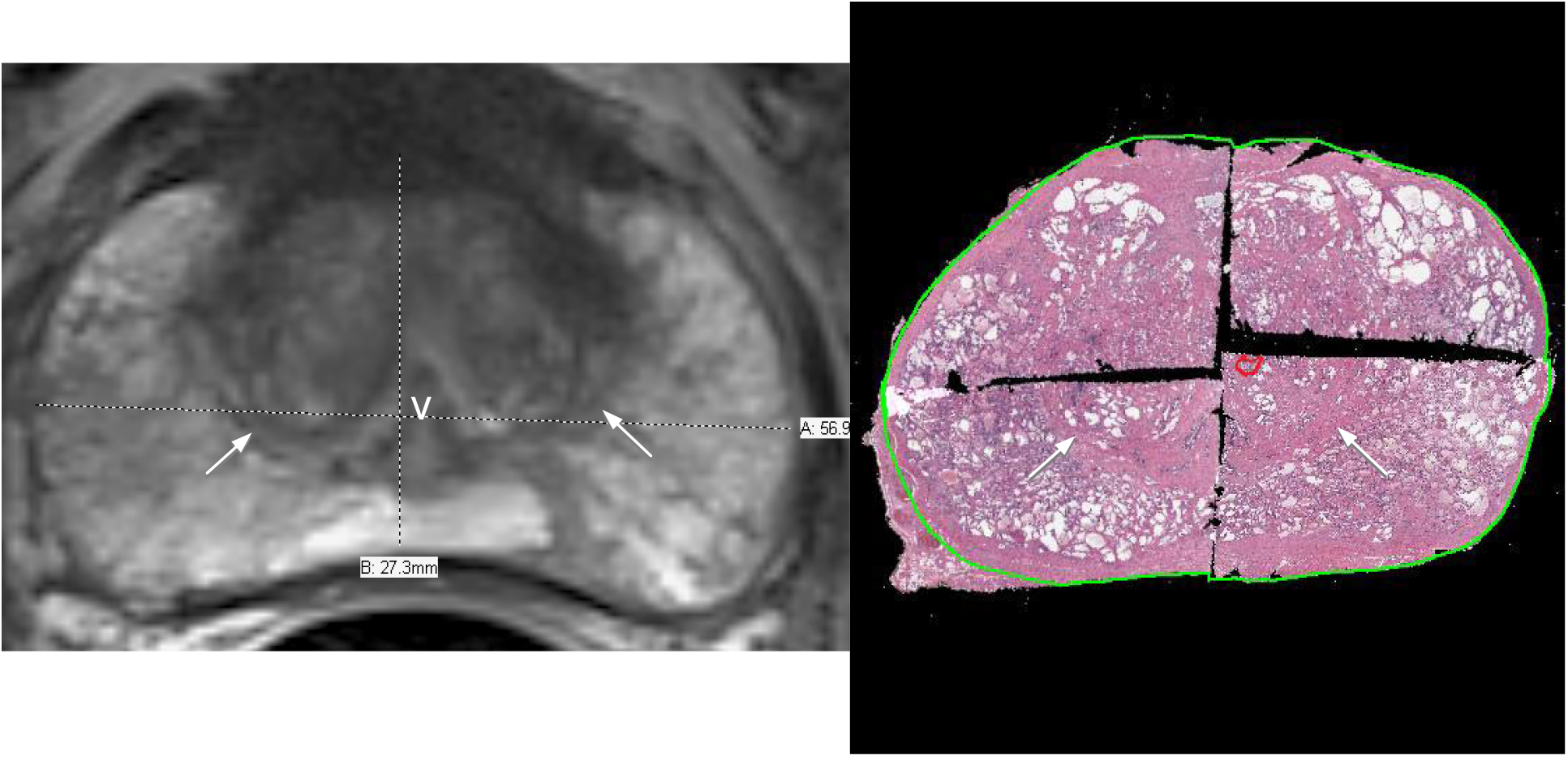
Axial T2-weighted MRI on left and corresponding axial H & E stained pathological specimen on the right. V=verumontanum. Arrows indicate surgical capsule boundary of the transition zone. Mean posterolateral peripheral zone thickness =16.2 mm in axial transverse plane.

**Fig 5.**
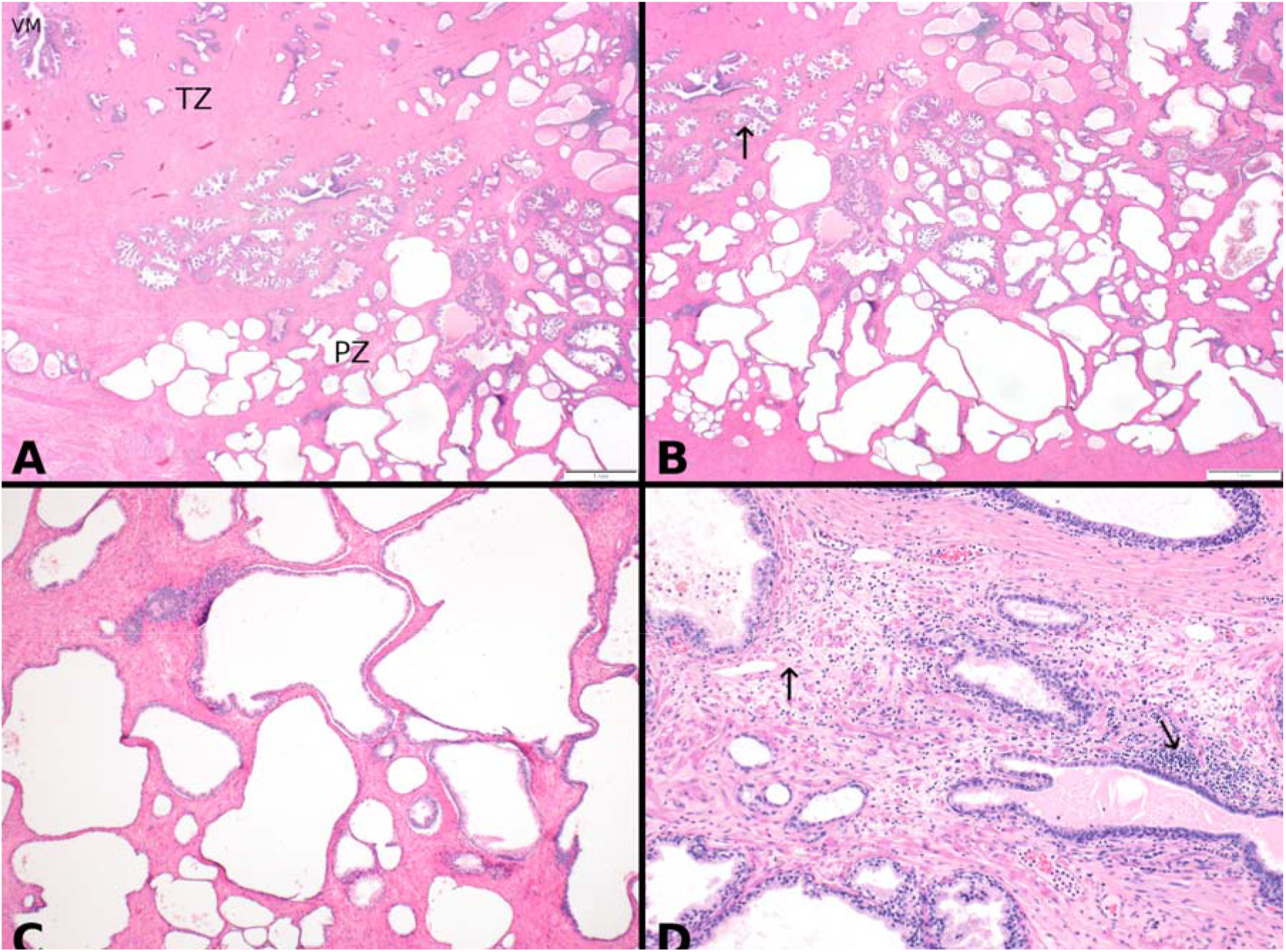
Hematoxylin and Eosin stained prostatectomy tissue sections, A showing peripheral zone (PZ) in lower left with verumontanum (VM), periurethral and transition zones (TZ) in upper right, B Distended and cystically dilated PZ glands in contrast to normal acini (arrow), C, higher magnification of cystically dilated PZ glands demonstrating flattened and atrophic single layered epithelial lining, D interstitial tissue between dilated glands showing chronic inflammation (right arrow) and edema (left arrow)

## Discussion

PZE is an extremely uncommon phenomenon, occurring in our practice an average of 3 times per year. Apart from the more common diagnostic findings of tumor or BPH, it is difficult to decide what to say if the PZ appears disproportionately thickened compared to overall prostate or TZ size. For this reason, we studied and reported the parameters of the normal-sized posterolateral PLPZ in patients with TPV ≤ 25 cc [4]. It was found that the mean PZ thickness was 10.0 mm with a standard deviation (SD) of 2.87 mm. We took 2 SD above the mean (5.74 mm) to represent the upper limits of normal. Thus, any posterolateral PZ measuring above 15.8 mm should be considered enlarged. Since there is ample evidence in the literature that the size of the PZ remains relatively stable throughout life [7, 8], this normal value can be extended to all age groups.

The 22 patients in our final cohort covered a wide range of TPV and TZV. Only 1 of 22 patients had a normal TPV (≤ 25 cc). It could reasonably be surmised that in several patients with increased to enormous TPV that the PZE is under-expressed in size due to compression. Mean TZV for 22 subjects was 20.6 cc, thus most had BPH. (Normal mean for TZ is 6.56 cc, 2SE equals 2.39-10.75) (4). There was no correlation between PLPZ and TZV (r=0.104, p= 0.630). No correlation was found between PLPZ thickness and PZV (r=0.236 p= 0.267). It is difficult to draw conclusions from PLPZ thickness as compared to AUASS since the data was incomplete.

It is important for the radiologist and urologist to recognize this condition in patients with over-all clinical or imaging prostatic enlargement who have little TZ enlargement on MRI but have obstructive symptoms. Even though the prostate may be big disproportionately due to PZE, the usual medical or surgical interventions may not be indicated or, at least, dynamic origin of the symptoms may be postulated, and treatment focused on alpha-adrenergic blockade. PZE is unlikely a contributor to obstruction.

TZI has been actively discussed in the urologic literature. Kaplan, et al [9], using TRUS, was first to suggest a strong correlation between TZI to patient symptoms and significant urodynamic abnormalities. TZI >0.50 has been reported to be associated with symptoms and signs of BPH [9, 10]. Some suggest a cut point of >0.40 as predictive of clinical BPH [11]. Others have not been able to demonstrate these associations [6, 12, 13].

Of the 22 patients in our series, only 3 had surgical resection of the entire prostate gland at our institution. We had convincing whole specimen pathology on only these 3 patients. 17 patients had needle biopsies. 7 were described as “negative or “benign”, 1 showed “atrophic glands and stroma”, 1 diagnosed as “BPH with acute and chronic prostatitis”, 3 had low grade adenocarcinoma, and 2 with high grade adenocarcinoma. No biopsy was requested for 3 patients. For the purposes of this paper, we did not attempt to correlate needle biopsy specimens for overall pattern due to limitations of biopsy sample size.

The overall pathological findings suggest a chronic inflammatory etiology for the finding of PZE. This was documented in the medical records of 4 of the 22 patients, but history of prostatitis was not found in the medical records of the 3 patients on whom we had prostatectomy tissue. However, because of the ubiquitous nature of prostatitis and possibility of incomplete patient historical memory, we can neither prove nor disprove a correlation with PZE. The absence of ductal dilation in the pathological material does not support the hypothesis of proximal ductal obstruction or injury of the verumontanum by instrumentation as causes of PZ glandular dilation. In fact, previous instrumentation did not prominently appear in our patient histories. Wahdera [14] postulated that acinar dilation may occur from chronic reflux of semen or urine secondary to pelvic muscle contraction. Nor was there evidence of ductal narrowing from scarring or chronic effects of prolonged contraction from drug effects [15].

A correlation between patients with metabolic syndrome and BPH has been suggested in the medical literature [16]. However, some doubt exists that diabetes mellitus represents an independent risk factor in development of prostatic enlargement [17]. It must be assumed that any effects on prostate size in diabetic patients are to be attributed to the TZ. There were 5 men (42%) in our study with history of type 2 diabetes mellitus, 2 of whom also had previous prostatitis. Therefore, diabetes and prostatitis cannot be excluded as having contributed to the inflammatory changes noted in the PZ of our pathological specimens.

An association between BPH and obesity has also been mentioned in the literature [18, 19]. The mean BMI in our patient cohort of 28.9 is in the normal range. Seven patients of 22 (32%) had a BMI ≥30 suggesting no correlation between PZ thickness and obesity.

There are some limitations to our study starting with measurement methods. Our selection of the largest axial transverse plane was arbitrary. In some patients, other axial or coronal planes may have different thicknesses that are more significant with respect to other measures, calculations, or clinical variables. Selection of exact PLPZ measurement boundaries and angles is subjective. Errors are most commonly due to misregistration of the boundaries at the prostatic apex and base in the axial planes. We attempted to mitigate this by making sure we were including these tissues in the coronal and sagittal views during manual contouring, but we can’t exclude the possibility of underestimating or overestimating volume. Measurement of TZI was biased by the conventional practice of including retrourethral (median lobe) tissue when calculating length in the ellipsoid formula for TPV. This would be expected to result overestimation of TPV which would falsely increase TZI, and PV would be expected to be falsely increased. We perseverated on this method as it is standard in the literature and allowed correlation with past studies. Other calculated values would likewise be affected. With only 3 optimal pathologic specimens to evaluate out of 22, these may not be representative of all cases. AUASS data was incomplete in the medical record.

## Conclusions

We describe a rare new MRI finding of PZ enlargement sometimes resulting in overall apparent increase in prostate size that must be distinguished from that caused by TZ enlargement when considering management decisions relating to obstructive symptoms. The pathologic findings of acinar dilatation and atrophy may, in some cases, be due to effects of chronic inflammation.

## Data Availability

Data is available for all reasonable requests.

